# Efficacy of Endovascular Thrombectomy in the Oldest Old (Stroke 90+ Study): A Community Center Experience

**DOI:** 10.1101/2023.03.26.23287556

**Authors:** Michael Brant-Zawadzki, Deborah Mastrolia, Junko Hara, Christopher Baker, Avinash Mesipam, Wallace Peck, David Brown

## Abstract

**Background:** While endovascular thrombectomy (EVT) is considered as the most effective treatment for a select group of patients with acute ischemic stroke and large vessel occlusion, its safety and efficacy in older patients are still debated especially in “read-world” settings. This study reports outcomes of EVT in acute ischemic stroke patients aged 90 and older in our community hospital setting.

**Methods:** Data between January 2018 and December 2022 were aggregated for all acute ischemic stroke patients with aged 90 and older at the time of EVT. Thirty-one patients met the criteria and were included in this report. The data valuables included, but are not limited to, demographics, stroke risk factors, thrombolysis in cerebral infarction (TICI), modified Rankin Scale (mRS), and NIH Stroke Scale (NIHSS).

**Results:** All 31 patients had improvement in TICI scale. One had symptomatic intracranial hemorrhage after EVT not related to the procedure, but likely on the basis of reperfusion breakthrough. Three patients expired prior to their discharge from non-stroke related causes. Of remaining 28, four expired, six went into hospice care, and four lost to follow-up by 30-days post DC. Of six hospice cases, one expired by 90-day post DC, and additional three were lost to follow-up. Given this data, 20/27 (74%) survived to 30 days and 16/24 (67%) to 90 days. For their NIHSS symptomatic categories, 15/28 (54%) patients improved, 10/28 (36%) remained the same, and 3/28 (11%) declined. For mRS, at 30-days post DC, 7/24 (30%) patients showed improvement, 7/24 (30%) remained the same, and 10/24 (40%) declined. At 90-days post DC, 7/21 (33%) showed improvement from DC, 4/21 (19%) remained the same, and 10/21 (48%) declined.

**Conclusions:** While a larger cohort study is necessary, our report supports the safety and efficacy of EVT in this patient aged 90 and older in a real-world setting.

## INTRODUCTION

Acute ischemic stroke is a leading cause of death and disability in United States affecting approximately 795,000 people each year [1]. Sudden-onset of weakness in an extremity or face, difficulty expressive or receptive aphasia, visual loss, confusion, and acute loss of balance or coordination are classic symptoms. Timely treatment of ischemic but not yet nonviable brain cells can prevent death and limits disability.

Over the past 20 years, intravenous thrombolysis (IVT) as proven treatment of acute ischemic stroke effects improvement in a relatively small percentage of strokes, ones mostly due to small vessel occlusions. Large vessel occlusions (LVOs) with their large clot burden are recalcitrant to IVT with suboptimal recanalization [2]. In fact, the implementation of IVT has been low internationally [3-5]. A recent series of well-designed and well-conducted randomized controlled trials (RCT) showed improved outcomes for LVOs in patients treated with endovascular thrombectomy (EVT) [6].

While EVT is considered as the most effective treatment for properly selected patients with acute ischemic stroke and large vessel occlusion by a recent editorial reviewed [6], its efficacy and appropriateness in older patients is still debated, as many studies have excluded patients over 80 years or included a much smaller sample size of such patients in the study. The landmark HERMES collaborative trial (Highly Effective Reperfusion Evaluated in Multiple Endovascular Stroke Trials), and its sub-study suggest not withholding EVT on the basis of age alone [7,8] although surveyed clinicians state that age still plays a key role when making individual thrombectomy decisions [9]. In addition, the most recent guidelines from the American Stroke Association/American Heart Association suggest more studies are needed for patients >=90 years especially in the “real-world setting” [10].

Therefore, we report “real world” outcomes of EVT in acute ischemic stroke patients 90 and older at the time of EVT. Our community hospital experience provides a demonstration of whether older patients may benefit from various stroke treatments outside an academic medical center setting. For this study, IRB exemption status approval was obtained from WCG IRB.

## METHODS

### Setting

Our community hospital, located in Orange County, CA, consists of two acute care campuses. The one in Newport Beach is designated by the Orange County Emergency Medical Services as a comprehensive stroke neurology receiving center with full time neuro-hospitalists backed by neuro-interventional radiology and neurosurgery. Newport Beach is also certified with DNV as a comprehensive stroke center and the other in Irvine is certified by DNV as a primary stroke center. In the most recent calendar year (2022), our emergency departments saw over 1,250 patients with stroke symptoms, making us the busiest stroke programs in Orange County, CA.

### Data

Electronic data records between January 2018 and December 2022 were aggregated without any personal identifiable and protected health information for the study. The inclusion criteria consisted of all acute ischemic stroke patients treated with EVT who were 90 years and older at the time of EVT. There were 31 patients who met the criteria and were included in this report.

The following variables were included:

- Demographics (i.e., age, gender, race)
- Stroke risk factors (i.e., BMI, hyperlipidemia, hypertension, history of stroke or TIA, atrial fibrillation, cancer, family history, coronary artery disease, myocardial infarction, diabetes, history, hypothyroid, peripheral vascular disease, deep vein thrombosis, and chronic obstructive pulmonary disease)
- Thrombolysis in cerebral infarction (TICI) scale at pre and post EVT
- Modified Rankin Scale (mRS) prior to stroke, at discharge (DC), and 30- and 90-days post DC
- NIH Stroke Scale (NIHSS) at admission and DC
- EVT relevant information (e.g., locations, admission time, discharge time)

### Analysis

Descriptive analyses were conducted to characterize the study population. EVT outcomes measured by TICI, mRS and NIHSS were analyzed. To determine factors that resulted in a more successful surgical outcome, linear regression models were created. These linear models used various demographic, LVO location, and risk factors as inputs, to ultimately output the change in measurement comparing TICI pre vs. post, mRS pre vs. 90 days post DC, and NIHSS admission vs. DC. Note that the TICI values were mapped to be on a numerical scale from 0-5 for the purposes of our analysis. From the linear regressions, coefficients for each of our input variables were generated. For mRS and NIHSS, given that higher values mean worse outcomes, a more negative coefficient for an input variable means that variable can reduce our outcome measurement, the change in mRS or NIHSS.

## RESULTS

**Table 1** shows sample characteristics. Among total 31 patients, average age was 92.9 +/-2.4 (min 90 and max 98). Twenty (65%) were female. Twenty-five (81%) were Caucasian and 5 (16%) Asian. Of all 31 patients, 25 (80.6%) had TICI scale 0 or 1 (no or minimum perfusion) at admission. After EVT, all patients had improvement in TICI scale (Table 1). Eleven patients received IV tPA treatment as well. One patient had a small symptomatic intracranial hemorrhage (SICH) a day after EVT not related to the procedure, and three patients expired prior to their discharge.

**Table 1.** Sample Characteristics

| Subject ID | Age Range | Gender | Race | BMI | Number of Risk Factors | LTSN | Door Time | Door to First Pass (min) | IV tPA | TICI |  | NIHS |  | mRS |  |  |  |
| --- | --- | --- | --- | --- | --- | --- | --- | --- | --- | --- | --- | --- | --- | --- | --- | --- | --- |
|  |  |  |  |  |  |  |  |  |  | Pre EVT | Post EVT | Admission | DC | Pre | DC | Post 30-Days | Post 90-Days |
| 1001 | 95-99 | F | Caucasian | 20.5 | 0 | 11:50 | 12:15 | 210 | Y | 0 | 2B | 17 | 23 | 0 | 5 | 6 | 6 |
| 1002 | 90-94 | F | Caucasian | 20 | 2 | 7:45 | 13:49 | 100 | - | 2A | 3 | 7 | 3 | 0 | 3 | 3 | 3 |
| 1003 | 95-99 | F | Caucasian | 22.2 | 3 | 13:50 | 14:32 | 138 | Y | 0 | 3 | 5 | 9 | 4 | 4 | 4 | 4 |
| 1004 | 90-94 | M | - | -99 | - | 9:15 | 10:08 | 138 | Y | 0 | 3 | 22 | exp | 0 | 6 | 6 | 6 |
| 1005 | 90-94 | F | Caucasian | 25.5 | 2 | 10:00 | 19:57 | 110 | - | 0 | 3 | 20 | 6 | 1 | 4 | 4 | 3 |
| 1006 | 90-94 | F | Caucasian | 28.4 | 3 | 23:00 | 0:13 | 95 | - | 0 | 3 | 20 | 6 | 1 | 4 | 4 | 4 |
| 1007 | 90-94 | F | Caucasian | 28 | 6 | 14:00 | 14:21 | 116 | - | 2B | 2C | 10 | 6 | 4 | 5 | 6 | 6 |
| 1008 | 90-94 | M | Asian | 20.34 | 1 | 15:00 | 17:15 | 149 | - | 0 | 3 | 13 | 13 | 4 | 4 | 6 | 6 |
| 1009 | 90-94 | F | Caucasian | 19.9 | 2 | 14:15 | 15:07 | 68 | Y | 0 | 2C | 28 | 23 | 3 | 5 | 6 | 6 |
| 1010 | 90-94 | F | Caucasian | 24.02 | 3 | 12:05 | 12:44 | 97 | - | 2A | 3 | 15 | 7 | 3 | 3 | hospice | 6 |
| 1011 | 90-94 | F | Caucasian | 18.7 | 3 | 12:00 | 12:27 | 58 | Y | 2B | 2C | 17 | 0 | 3 | 4 | 3 | 3 |
| 1012 | 90-94 | F | Caucasian | 14.2 | 3 | 19:30 | 8:45 | 90 | - | 0 | 3 | 20 | 23 | 4 | 5 | hospice | hospice |
| 1013 | 90-94 | F | Caucasian | 24.8 | 4 | 11:15 | 11:42 | 82 | Y | 1 | 3 | 42 | 2 | 1 | 4 | 3 | 2 |
| 1014 | 95-99 | F | Caucasian | 22.4 | 2 | 18:40 | 18:40 | 150 | - | 0 | 2C | 24 | 3 | 3 | 4 | 4 | 4 |
| 1015 | 90-94 | F | Caucasian | 19 | 1 | 17:00 | 8:40 | 66 | - | 2A | 3 | 8 | 6 | 0 | 4 | 1 | 1 |
| 1016 | 95-99 | F | Caucasian | 27.88 | 2 | 18:30 | 19:29 | 117 | Y | 0 | 3 | 31 | 8 | 3 | 4 | 3 | 3 |
| 1017 | 90-94 | M | Caucasian | 23.41 | 2 | 17:08 | 17:46 | 136 | Y | 0 | 2B | 35 | 30 | 0 | 5 | - | - |
| 1018 | 95-99 | M | Asian | 23.57 | 2 | 8:45 | 9:32 | 67 | Y | 2A | 2C | 21 | 7 | 0 | 4 | 3 | 3 |
| 1019 | 90-94 | M | Caucasian | 24.11 | 3 | 15:30 | 21:36 | 104 | - | 0 | 2B | 20 | 20 | 0 | 5 | hospice | hospice |
| 1020 | 95-99 | F | Asian | 28.28 | 5 | 20:30 | 21:24 | 140 | Y | 1 | 3 | 25 | 0 | 3 | 3 | - | - |
| 1021 | 90-94 | M | Asian | 33.39 | 2 | 23:00 | 7:43 | 112 | - | 1 | 3 | 16 | 6 | 0 | 4 | 2 | - |
| 1022 | 90-94 | F | Caucasian | 20.62 | 4 | - | 10:59 | 131 | - | 0 | 3 | 13 | exp | 0 | 6 | 6 | 6 |
| 1023 | 95-99 | F | Asian | 21.96 | 4 | 18:00 | 18:52 | 90 | - | 0 | 3 | 33 | 14 | 4 | 4 | 4 | - |
| 1024 | 90-94 | F | Caucasian | 17.87 | 2 | 7:00 | 12:58 | 70 | - | 0 | 3 | 13 | 14 | 0 | 5 | 3 | - |
| 1025 | 90-94 | M | Caucasian | 21.85 | 3 | 16:45 | 17:24 | 97 | Y | 1 | 2C | 17 | 10 | 0 | 5 | hospice | hospice |
| 1026* | 90-94 | M | Caucasian | 33.25 | 7 | 1:00 | 9:07 | 88 | - | 0 | 3 | 6 | 1 | 0 | 4 | 4 | 1 |
| 1027 | 90-94 | M | Caucasian | 24.37 | 3 | - | 18:57 | 342 | - | 0 | 2B | 5 | 6 | 3 | 4 | hospice | hospice |
| 1028 | 90-94 | F | Caucasian | 18.13 | 0 | 5:00 | 11:38 | 83 | - | 0 | 2C | 18 | 26 | 0 | 5 | hospice | hospice |
| 1029 | 90-94 | M | Caucasian | 23.65 | 3 | 13:30 | 22:50 | 125 | - | 0 | 2C | 29 | 17 | 2 | 5 | - | - |
| 1030 | 90-94 | F | Caucasian | 22.31 | 5 | 6:00 | 8:37 | 62 | - | 0 | 3 | 27 | 12 | 4 | 5 | - | - |
| 1031 | 90-94 | M | Caucasian | 24.77 | 4 | 20:18 | 20:48 | 118 | - | 0 | 2C | 20 | exp | 0 | 6 | 6 | 6 |

\*: Symptomatic intracranial hemorrhage; LTSN: last time seen normal; DC: discharge; NIHS: NIH stroke scale; mRS: modified Rankin scale; TICI: thrombolysis in cerebral infarction scale; exp: expired; -: missing data or lost to follow-up.

Of remaining 28, four expired, six went into hospice care, and four lost to follow-up by 30-days post DC. Of six hospice cases, one expired by 90-day post DC, and additional three were lost to follow-up. Thus, among patients with available data, 20/27 (74%) survived to 30 days and 16/24 (67%) to 90 days.

**Figure 1** compares the number of patients in each NIHSS symptomatic category at admission vs post-DC. Three patients who expired prior to DC were excluded. For their symptomatic categories, 15/28 (54%) patients improved, 10/28 (36%) remained the same, and 3/28 (11%) declined.

**Figure 1.**
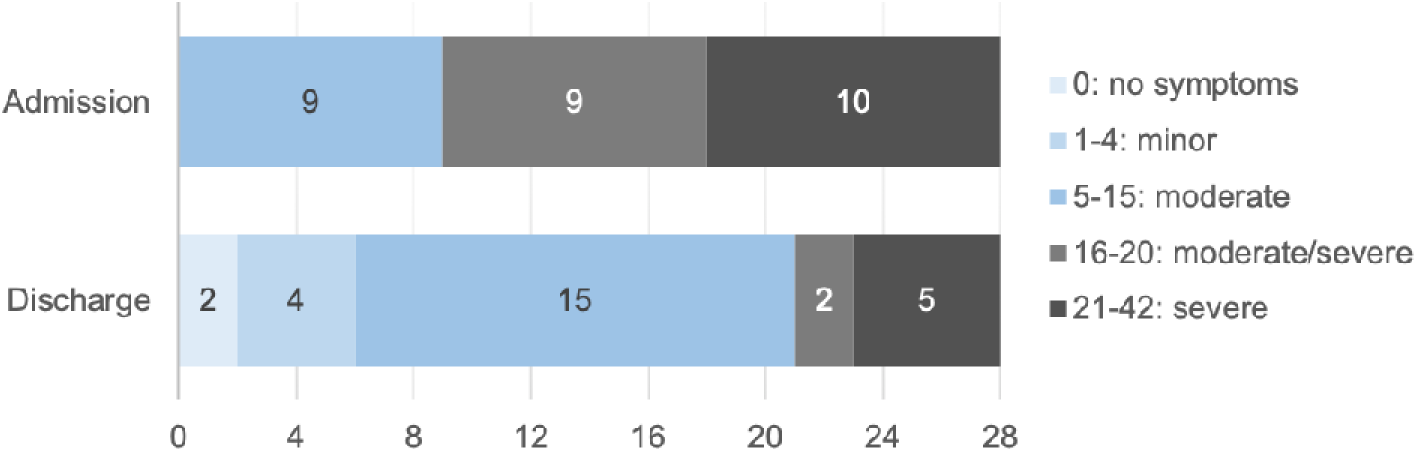
Number of Patients per NIH Stroke Scale at Admission vs. Post-DC.

**Table 2** shows the number of patients in each mRS at DC, 30- and 90-days post DC. By 30-days post DC, data were available for total 24 patients, among which 4 expired and 6 went into hospice care. Of those 10, seven were severe disabled stage (mRS 5) at DC. By 90-days post DC, data were available for total 21 patients, among which 5 expired and 5 remained at hospice care.

**Table 2.** Number of Patients per mRS Group at 30-Days vs. 90-Days Post-DC.

|  | mRS |  |  |  |  |  |  | hospice | lost to follow-up |
| --- | --- | --- | --- | --- | --- | --- | --- | --- | --- |
|  | 0 | 1 | 2 | 3 | 4 | 5 | 6 |  |  |
| DC | 0 | 0 | 0 | 3 | 14 | 11 | 0 | 0 | 0 |
| 30 Day DC | 0 | 1 | 1 | 6 | 6 | 0 | 4 | 6 | 4 |
| 90 Day DC | 0 | 2 | 1 | 5 | 3 | 0 | 5 | 5 | 7 |

**Figure 2** shows changes in mRS at 30- and 90-days post DC. At 30-days post DC, 7/24 (30%) patients showed improvement, 7/24 (30%) remained the same, and 10/24 (40%) declined. At 90-days post DC, 7/21 (33%) showed improvement from DC, 4/21 (19%) remained the same, and 10/21 (48%) declined.

**Figure 2.**
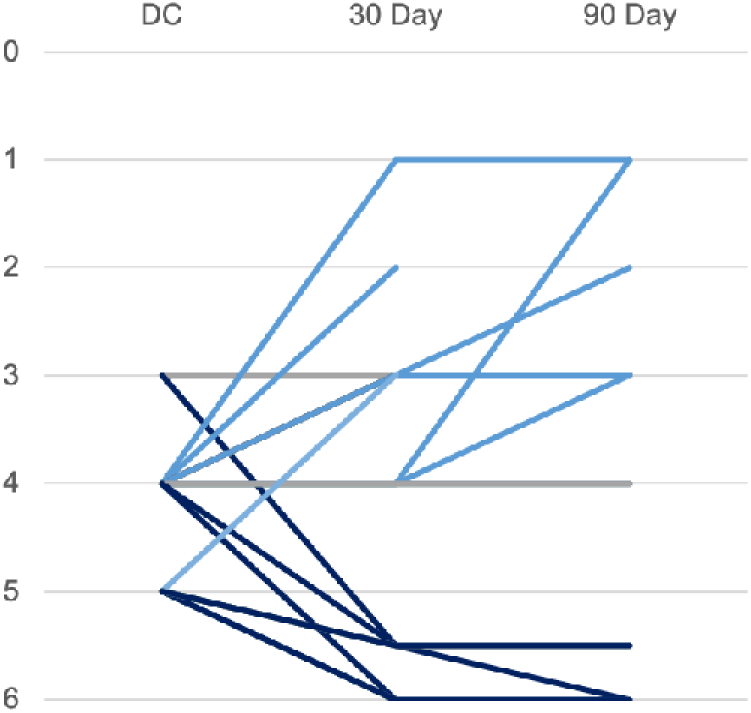
Change in mRS at 30-Days vs. 90-Days Post-DC. Light blue line indicates mRS improvement, gray line no change, and dark blue decline during the course of 90-days follow up compared to DC.

Due to the small sample size, two separate approaches were taken. The first approach was creating a linear model using the following variables: age, race, BMI, the count of all risk factors, door to first pass (in minutes), IV tPA administration, and LVO locations. The second approach to the linear regressions involved the following valuables: age, race, BMI, specific risk factors (i.e., Afib, HLD, HTN), the count of all other risk factors, door to first pass (in minutes), IV tPA administration, and LVO locations. All of these coefficients reflect the predicted relationship between input variables and the outcome. However, none showed statistically significant results, indicating that the input variables did not significantly predict the outcomes in this sample.

## DISCUSSION

This report has documented safety and benefit of EVT in patients who are 90 years and older in a community hospital setting, further supporting EVT in eligible advanced aged patients. Our outcomes were equivalent to, although there are few, other published studies [10].

In our cohort, there was one patient (patient ID: 1026) with a small SICH after EVT showing NIHSS decline from 6 to 17. However, his NIHSS improved to 1 at DC. This SICH is most likely related to reperfusion breakthrough as the patient had basilar thrombosis upon arrival and had 2 × 1 cm hematomas in left basal ganglia with mild mass effect on the internal capsule. The patient was also administered IV tPA per our protocol prior to EVT.

Three patients expired prior to their discharge. The first patient (ID: 1004) had LICA terminus occlusion with successful EVT outcomes. However, he had LMCA edema and brain herniation and expired 3 days after EVT. The second patient (ID: 1022) had left PICA with successful EVT with residual basilar thrombus and dissection of basilar artery. She did not wake up after EVT, remained intubated on ventilator, and expired four days later. The third patient (ID: 1031) had LMCA with successful EVT. However, he had multiple comorbidities including advanced Parkinson’s disease, and showed no neurologic improvement, continued to have aphasia and dysphagia inability to control secretions leading to aspiration pneumonia. He expired 18 days after EVT.

Among patients with available follow-up data, mortality rates were 26% and 33% at 30-days and 90-days post DC, respectively. Some published studies report mortality rate ranging from 31 to 37% for patients who are 80 years and older with EVT [11-13] although this rate is higher than those for younger patients. McDonough et al. compared mortality rate for patients >=85 years old demonstrating better mortality rate of 31% with EVT compared to 54.3% for those received standard of care [11].

In our cohort, all patients showed improvement in perfusion rate in TICI scale (Table 1). NIHSS also showed improvement with 63% reduction in mandate/severe to severe (NIHSS 16-42) stages (Figure 1). Although only 2 and 3 patients shifting its score to 0-2 at 30-days or 90-days post DC, respectively, mRS showed improvement in symptomatic categories (Figure 2) in 30% and 33% patients at 30-days and 90-days post DC, respectively, further supporting EVT in advanced aged adults.

While not the purpose of this study, a legitimate discussion regarding the cost benefit of treating LVO related acute stroke on a quality of life years (QUALY) basis can be in this advanced age population would have to include the cost of long term care for survivors, Also, LVO of distal carotid and middle cerebral artery trunks leads to mortality in the majority of patients prior to the availability of EVT [14,15], thus death impacts a cost per life year saved analysis in the oldest old.

A recent US nationwide retrospective cohort study using the Get with the Guidelines–Stroke (GWTG-Stroke) program [16] demonstrates steady increase in the use of EVT in patients aged 80 years and older from 3.3% in quarter 3 of 2012 to 20.8% in quarter 2 of 2019 in the study cohort (n = 12,768). This trend was similar for patients <80 (n = 29,654) during the same time period. However, the EVT treatment rates still remain lower than in younger patients. With more EVT outcome data especially from real-word setting, the use of EVT can be further evaluated in this cohort.

The limitation of this study is the small sample size. While this case series demonstrates favorable EVT outcome for this oldest old population, the outcome should be viewed with its small sample size in mind. However, this report warrants further studies in this cohort.

## Data Availability

Portion of the data used for outcome analyses are included as Table 1. Additional data is available upon request.

https://figshare.com/

## ACKNOWLEDGMENTS

We thank the multidisciplinary stroke team at Hoag for their dedication and tireless work to provide the best possible outcomes for our patients. This includes the Emergency Department, Neuro Interventional Radiology, Neuroscience Intensive Care Unit, 3 East Advanced Brain and Spine Unit and Neuro Rehabilitation Services. We thank our Stroke Nurse Navigator, Victoria Tomczak for her data assistance. We also acknowledge our neurology team, Drs. Andrew Ly, Jason Muir, Jose Puancgo, David Millet, Victor Doan, James Park, and Kaveh Saremi, as well as our interventional radiology technologists, George Roth, Kari Malmberg, Jon Hanaoka, Sam Gentry, Kyle Higashida, Naomi Yoshizawa, and Farina Usman, for their support on stroke treatment.

